# Multi-lineage natural gene therapy mediated by embryonic triploid mosaicism in the context of Fanconi anaemia

**DOI:** 10.1101/2025.10.29.25337140

**Authors:** Michael Sharp, Caitlin Harris, Chirantani Mukherjee, Stevan Novakovic, Elissah Granger, Roser Pujol, Gerard Muñoz-Pujol, Elva Shi, Karen Dun, Cesar Salinas-La Rosa, Zhen Hou Xu, Mark D. Pertile, Krystle Standen, Rebecca Walsh, Andrew J. Deans, Eunike Velleuer-Carlberg, Jonathan Moses, Adayapalam Nandini, Adam Nelson, Lisa Worgan, Jordi Surrallés, Wayne Crismani

**Author notes:** Co-first authors (Michael Sharp, Caitlin Harris).

## Abstract

Fanconi anemia is a rare inherited bone marrow failure syndrome caused by inactivation of genes in the Fanconi anemia/BRCA DNA repair pathway. We report a patient with X-linked Fanconi anemia, and atypical physical features whose genetic diagnosis was initially inconclusive. Over time, his bone marrow karyotype shifted from diploid (46,XY) to triploid (69,XXY). The triploid cells lacked the Fanconi anemia cellular phenotype, enabling sustained hematopoiesis and providing an unexpected route to phenotypic rescue. Genomic analysis indicated early post-zygotic incorporation of the second polar body as the triploid origin. These findings suggest that the selective advantage of restored DNA repair in hematopoietic stem cells, outweigh the potentially deleterious effects of triploidy.

## Introduction

Fanconi anemia (FA) is a rare inherited condition which predisposes to progressive bone marrow failure, early-onset cancers, and multiple congenital anomalies with substantial impact on quality of life^1–3^. FA is caused by inactivation of genes in the FA/BRCA DNA repair pathway, most often autosomal recessive^4^. One subtype is X-linked and caused by loss of function variants in *FANCB* (HGNC:3583)^5^. All subtypes have a genomic instability phenotype. FA is diagnosed with a chromosome fragility assay where crosslinking drugs diepoxybutane (DEB) or mitomycin C (MMC) are used to exacerbate the underlying DNA repair defect in FA patient cells^6,7^. Mosaicism for restored DNA repair in FA is well described; however, it is usually somatic. Somatic mosaicism in the context of FA, typically refers to advantageous secondary or reversion mutations in a hematopoietic stem cell (HSC) that restores partial or complete function of the relevant FA gene, which leads to a proliferative advantage of the HSC and its daughter cells ^8,9^, and is associated with improved hematology in FA^10^. The proliferative advantage of gene-corrected HSCs underlies the rationale for hematopoietic gene therapy without genotoxic conditioning^11^. Here we describe a case of embryonic triploid mosaicism of maternal origin which rescues hematopoiesis and the underlying DNA repair defect in FA.

Triploidy is not compatible with *ex utero* human life. Most triploid pregnancies result in early miscarriage^12^. Rare cases that proceed to full term typically do not survive more than a few days. Embryonic triploid mosaicism is a rare condition where some cells in the body are triploid, and some are diploid. Triploid mosaicism has a highly variable presentation and is likely underdiagnosed^13,14^.Triploid cells are more frequently observed in muscle biopsies with only 5% of cases having triploidy in the hematopoietic system^14^. The origin of full triploidy or triploid mosaicism can be either from the father or from the mother, with parent-of-origin effects^15^. Here we describe a young boy who has a diagnosis of X-linked FA, caused by disruption of *FANCB*. In this patient, we observed a progressive expansion of a triploid (69,XXY) hematopoietic lineage carrying a functional maternal *FANCB* allele (Fig. 1). These triploid cells lacked the FA cellular phenotype and now predominate in the bone marrow. We describe the clinical and molecular features of the patient and provide evidence that embryonic triploid mosaicism has enabled functional hematopoiesis, suggesting an unexpected route to phenotypic rescue.

**Figure 1.**
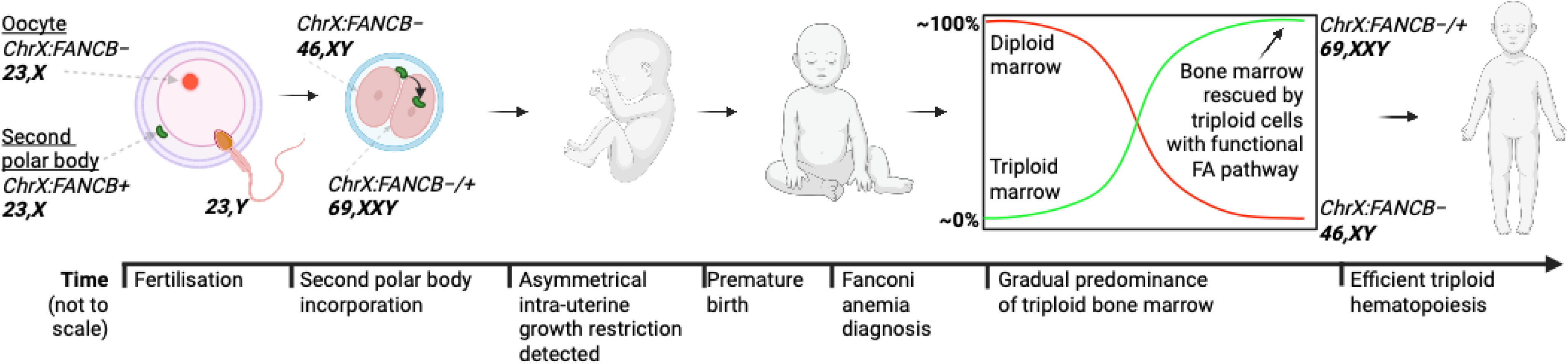
Embryonic triploid mosaicism in the context of FA is associated with a progressive shift to triploid bone marrow. Fertilization resulted in a diploid (46,XY) zygote that lacked *FANCB* function. Incorporation of the second polar body, with a functional *FANCB*, at the post-zygotic stage results in a mosaic embryo (46,XY, lacking *FANCB* function, and 69,XXY, with one functional *FANCB* allele). Over time the bone marrow and peripheral blood become predominantly triploid and are associated with improved complete blood counts.

## Results

### Clinical presentation and initial genetic evaluation

A young boy male was evaluated by paediatrics and genetics given multiple minor dysmorphic features were detected. A maternal family history of FA, consistent with X-linked inheritance, prompted targeted genetic evaluation (not shown). Chromosome fragility tests confirmed the diagnosis of FA.

### Functional rescue confirms FANCB as the causal gene

Given the X-linked inheritance pattern of FA in this family, *FANCB*^5^ was considered the most likely causative gene. To test this hypothesis, we performed complementation studies using lentiviral transduction of *FANCB*. Immortalized fibroblasts were established from the left and right sides of the body – given the physical asymmetry – and a lymphoblastoid cell line derived from peripheral blood. Chromosome breakage testing confirmed the FA diagnosis in both fibroblast cultures, consistent with previous findings in peripheral T-cells. Therefore, we chose to focus on characterising left-side fibroblasts. Genetic complementation was achieved in the left-side fibroblasts, with lenti-viral transduction of *FANCB*, rescuing chromosome fragility at the highest DEB concentration tested (Fig. 2A, Sup. Table 1, DEB 0.05 µg/mL; Fisher’s exact test, Haldane-Anscombe–corrected odds ratio 70, 95% CI 10–500, p < 0.001). In addition, *FANCB* transduction restored FANCD2 focus formation (Fig. 2B, Kruskal–Wallis test followed by Dunn’s multiple comparisons, all adjusted p < 0.05). *FANCB* transduction also had increased MMC resistance in the patient’s fibroblasts. Although Welch’s t-tests did not reach statistical significance given the small number of technical replicates (experiment 1: t = 2.42, df = 2.0, p = 0.14; experiment 2: t = 2.78, df = 2.0, p = 0.11), the consistent increase across independent transductions supports functional rescue (Fig. 2C), further supporting rescue of FA pathway function. These results support *FANCB* as the causal gene underlying the FA phenotype in this patient.

**Figure 2.**
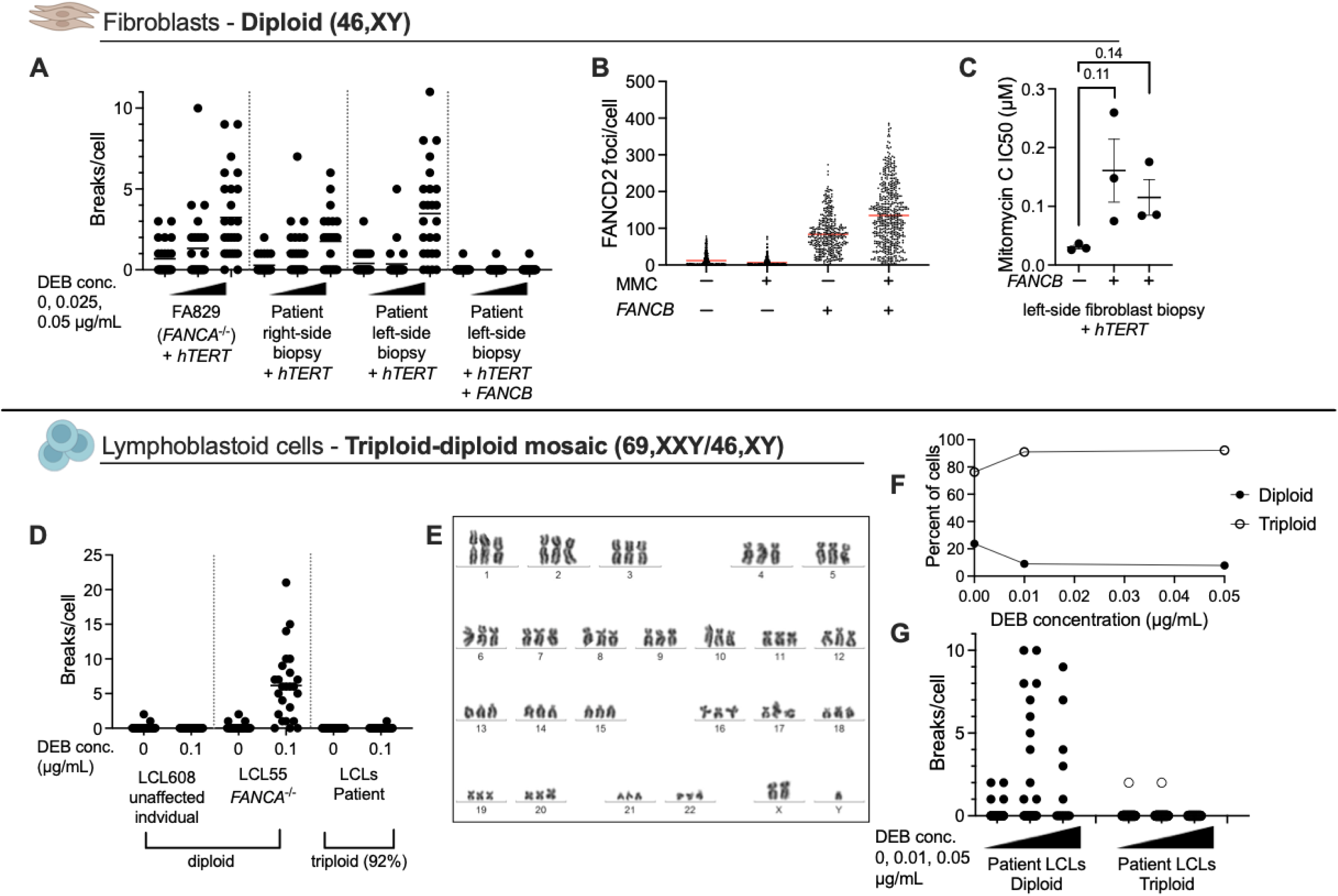
Triploid mosaicism is associated with *FANCB-*dependent rescue of the FA phenotype. **A)** Patient fibroblasts test positive to FA but chromosome fragility can be rescued by lentiviral transduction of *FANCB*. **B)** Patient fibroblasts transduced with *FANCB* rescues FANCD2 focus formation. **C)** *FANCB* transduction rescues mitomycin C sensitivity in patient fibroblasts. Exp1 and Exp2 represent independent transductions with FANCB. **D)** Patient LCLs do not have a chromosome fragility phenotype and have a high proportion of triploid cells. **E)** LCLs continuously cultured had a predominantly triploid karyotype (69,XXY), with a minority being diploid (46,XY). **F)** The proportion of diploid cells the patient’s LCLs decreases with increasing doses of DEB. **G)** LCLs Chromosome fragility testing reveals two populations of cells in the patient LCLs; diploid cells sensitive to mitomycin C and triploid cells resistant cells. For panels A, D, G statistical analysis was performed on the number of aberrant cells. Plots show the number of chromatid per cell for visual clarity in A, D and G.

Long-read RNA sequencing of the patient’s fibroblasts revealed mis-splicing of the *FANCB* transcript, resulting in the exonization of a portion of a predicted transposon (Sup. Fig. 1). Targeted cDNA sequencing confirmed the inclusion of an additional 110 nucleotides from a retained intronic region (Sup. Fig. 2). Within this region we identified a single-nucleotide deletion, *FANCB* (NM_001018113.3:c.1326+594del), that causes a frameshift and introduced a premature stop codon (Sup. Fig. 1). This variant was absent from gnomAD and ClinVar.

Given that *in silico* models (SpliceAI) also predicted this intron retention and that the variant segregated with the phenotype (not shown), we prioritized further analysis of the variant. A minigene assay comparing the patient and control sequences demonstrated that this variant is sufficient to cause the observed intron retention in *FANCB* and therefore the introduction of a premature stop codon (Sup. Fig. 3). Based on these combined lines of evidence – including segregation, absence in population databases, splicing predictions, and functional validation – we consider this variant as pathogenic (PP1, PM2, PVS1, according to ACMG criteria^16^).

Triploid cells predominate in hematopoiesis and lack the FA phenotype

Chromosome fragility studies with lymphoblastoid cells (LCLs) from the patient tested negative for FA at the highest DEB concentration (Fig. 2D, Sup. Table 2, DEB 0.1 µg/mL; Fisher’s exact test, Haldane-Anscombe–corrected odds ratio 0.8, 95% CI 0.2–3.3, p > 0.99). We also observed that 92% of the metaphase cells were triploid, with a 69,XXY karyotype (Fig. 2E). We tested an earlier passage, which contained a higher proportion of diploid cells and found that diploid LCLs were sensitive to DEB-induced chromosomal aberrations, whereas the triploid LCL lineage was not sensitive at the highest concentration tested (Fig. 2F, 2G, DEB, 0.05 µg/mL; Fisher’s exact test, Haldane-Anscombe-corrected odds ratio 51.1, 95% CI 2.6-1015, p < 0.001). The patient’s immortalized fibroblasts also had a low proportion of triploid cells (left side fibroblasts, 3.5%; 3/88, right side fibroblasts, 4.1% 5/127). Therefore, we investigated if the triploid karyotype was also present *in vivo* in the patient. Rescanning the slides from the chromosome fragility test – which predominantly analyzes stimulated T cells – at the time of diagnosis revealed 0.3% triploid cells (n=628) from the no DEB condition. We revised cytogenetic karyotyping reports from routine bone marrow aspirates. We found that there had been previous cytogenetic evidence of a limited number of triploid cells in the bone marrow at which started to emerge after diagnosis of FA (1%; 2/169) and with increasing proportions of triploid cells at the next bone marrow aspirate (10%; 11/110). At the next two bone marrow aspirate the proportion of triploid cells, 69,XXY, had increased to 95% (104/110) and 100% (215/215, Sup. Fig. 4) respectively.

### Triploid mosaicism is maternal in origin and arises post-zygotically via second polar body incorporation

To determine if the triploid lineage was confined to hematopoietic tissue or more widely distributed, we analyzed buccal epithelium isolated with an Orcellex Brush and cells from the urine. Analysis of the left and right buccal epithelium revealed a high proportion of 69,XXY cells (34% and 87% respectively), consistent with triploid mosaicism of embryonic origin. A pathologist confirmed that a sufficient proportion of the buccal cells analyzed were epithelial, ensuring that the triploid lineage was present in non-hematopoietic tissues. Similarly, triploid cells were detected in the urine (40%, Sup. Fig. 5). The high proportion of triploid cells in the buccal epithelium is consistent with a competitive advantage conferred to cells with a functional FA pathway, similar to what was observed in the hematopoietic system.

Whole-genome sequencing of predominantly triploid LCLs and predominantly diploid fibroblasts identified ∼43,000 variants unique to the triploid lineage. These variants matched maternal alleles, indicating a maternal origin of the additional chromosome set. Genome-wide analysis of these postnatal samples showed maternal alleles to be homozygous near centromeres and a tendency towards heterozygosity near telomeres (Fig. 3A, Sup. text), consistent with crossing over distributions during meiosis I followed by incorporation of the second polar body. On reanalysis of the prenatal amniocentesis SNP microarray data, we identified the same maternal inheritance at low levels (Fig. 3B, Sup. Fig. 6), consistent with the findings from postnatal samples. Extending this analysis of genomic sequencing samples from the patient revealed an increase in triploidy in the hematopoietic system over time, where it was undetectable in peripheral blood at the first collection time point but predominant in peripheral blood and the bone marrow at later time points (Fig. 3C, Sup. Fig. 7, Sup. text).

**Figure 3.**
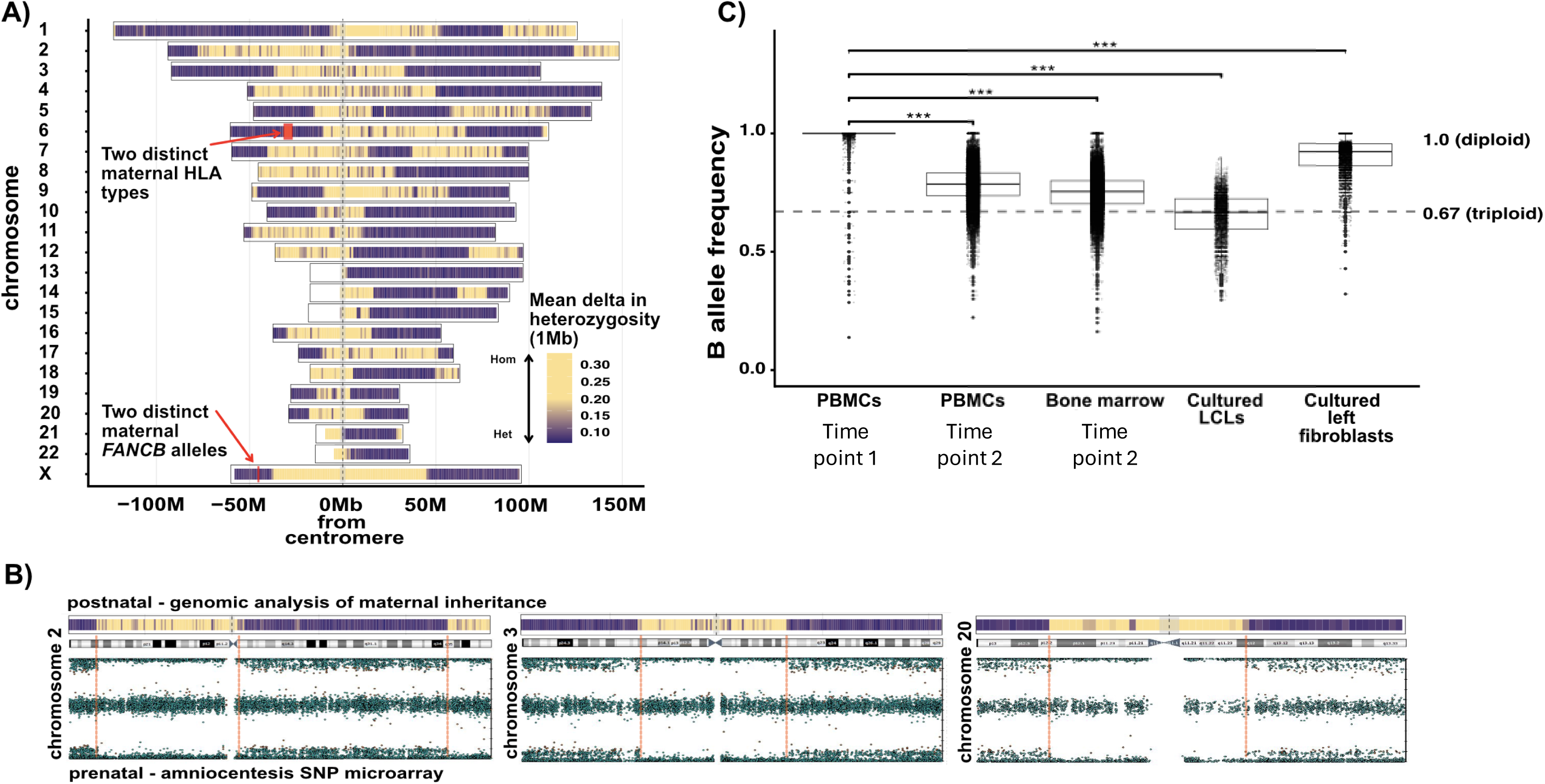
An extra set of chromosomes was inherited from the same maternal meiosis. A) Chromosomes are aligned by their centromeres. Patient variants unique to the triploid lineage are considered and binned in 1 Mb regions and are colored according to whether their maternal inheritance is heterozygous (purple) or deviates from the expected allele frequency (increased yellow) and towards homozygosity. B) A comparison of prenatal amniocentesis SNP microarray data (lower) and genomic analysis from panel A (upper). Dashed vertical red lines highlight sites of maternal meiotic recombination which are concordant between the two different approaches. Three representative chromosomes are shown and are not to scale. All chromosomes are shown in Supplementary Figure 6. C) Hematopoietic samples show increasing genome-wide triploid allele frequencies at the most recent routine collection compared to the time of diagnosis.

Orthogonal support for triploid lineage expansion came from longitudinal HLA typing performed as part of anticipatory planning for potential future hematopoietic stem cell transplantation. DNA was first extracted from peripheral blood collected at around the time of diagnosis for HLA typing. Sequencing results were initially assessed as unremarkable and gave a clear typing consistent with inheritance of one copy of chromosome 6 from each of the patient’s parents using high resolution sequencing. Subsequent reanalysis of the data using relaxed parameters revealed low read counts which supported the presence of an additional set of *HLA* alleles for the other maternal haplotype, consistent with a minor triploid population at that time. DNA extracted from peripheral blood at at a later age revealed high levels of three *HLA* alleles at all tested loci (HLA-A, B, C, DRB1, DRB3/4/5, DQA1, DQB1, DPA1 and DPB1). The previously detected paternal haplotype was still present but now both maternal haplotypes appeared to be present with high representation. This would be consistent with the expansion of a maternally derived triploid lineage, consistent with the genome-wide analyses (Fig 3A, Sup. Fig. 6).

Due to the predominance of triploid hematopoiesis associated with a functional FA pathway, peripheral blood counts remain relatively stable and within normal ranges (Sup. Fig. 8). The most recent marrow aspirate was moderately hypocellular for age with mildly reduced granulopoiesis, megakaryopoiesis and mild dyserythropoiesis consistent with FA. Longer follow-up will be needed to determine if triploid hematopoiesis can be stably maintained.

## Discussion

We report on a case of triploid mosaicism in the context of X-linked (*FANCB-*deficient) Fanconi anemia. The triploid lineage carries a functional maternal copy of *FANCB*, which provides functional rescue. The bone marrow was initially diploid, 46,XY, with lack of *FANCB* function. However, over time the triploid lineage, 69,XXY, with a functional *FANCB* and FA DNA repair pathway became the predominant lineage in the bone marrow and the peripheral blood. In this context, triploid hematopoiesis appears to be efficient and clinically competent. The duration of follow-up is a limitation, and ongoing observation will be required to assess long-term stability of triploid hematopoiesis and genomic stability.

Moreover, atypical karyotypes could predispose to leukemia or other associated diseases.

Whole-genome analysis revealed that both maternal sets of chromosomes in the triploid lineage originated from the same meiotic event. This is evidenced by the retention of maternal homozygosity at the centromeres and proximal genomic regions, consistent with post-zygotic incorporation of the second polar body. Further, the finding that all 46 maternal chromosomes originate from a single meiosis argues against an alternative hypothesis of embryo-embryo fusion as the source of the triploid mosaicism. The asymmetrical IUGR and placental insufficiency associated with low fetal fraction of cfDNA are also consistent with previous reports that the parental origin of an extra set of chromosomes affect these specific fetal and placental phenotypes^15^. The clinical presentation with dysmorphic features, asymmetry of growth and skin pigmentation could also be explained by the triploid mosaicism.

The competitive growth advantage conferred by a functional FA pathway in the triploid cells likely was the cause of the triploid clone expansion. While other factors could theoretically contribute to the selective advantage of the triploid lineage, we consider this unlikely for two reasons: 1) It is well documented that somatic reversion of hematopoietic stem cells leads to a competitive growth advantage and this embryonic form of genetic rescue is somewhat akin to that rescue mechanism^9^; 2) other cases of triploid mosaicism – without the context of FA – have not reported such a marked shift from nearly undetectable to complete triploid marrow, and the expansion observed here is most plausibly attributed to functional rescue *via FANCB*. In the context of this patient, we hypothesise that the benefits of functional DNA repair might outweigh the disadvantages of triploid haematopoiesis. These observations raise questions about whether other disorders with a selective growth disadvantage might occasionally be mitigated through similar forms of embryonic mosaicism which could mediate natural gene therapy, underscoring the importance of vigilance in interpreting unusual karyotypes.

## Data Availability

All data produced in the present study are available upon reasonable request to the authors

## Acknowledgements

We thank the patient, his parents and the extended family for their willingness to participate in this research. Thank you to Marta Cifuentes Ochoa, Irene Gallego Romero, Davis McCarthy, Mark Cigan and Ludmila Matyakhina for helpful discussions. Thank you to Lucy Perez Kempster and MCRI for assistance with the production of the LCLs, and to The cytogenetics team at Pathology Queensland. Figure 1 and parts of Figure 2 were created with Biorender.com.

## Supplementary text

*The maternal origin of triploid mosaicism –* To determine the origin of the extra genome in the patient, we identified variants in the triploid lymphoblastoid genome and diploid fibroblast genome. The non-reference variants which were unique to the triploid LCLs were maternal in origin, with 43,337 variants coming from the mother and 128 from the father, after having filtered out repetitive regions and selecting high quality variants.

*Triploidy arose via incorporation of the second polar body (Fig. 3A) –* To analyse the genome-wide maternal inheritance patterns of the extra genome in the triploid lineage of the patient, we performed the following analysis.

1. We filtered for variants which were heterozygous in the mother’s germline and homozygous reference or homozygous alternate in the father’s germline. We refer to these variants as “triploid-informing-variants” below.
2. Using a VCF from the LCLs filtered for triploid-informing-variants, and allele frequencies were calculated based on the proportion of reads supporting the B allele frequency (BAF). With the understanding that the LCLs were mostly triploid, the triploid-informing-variants from step 1 (above) with BAFs at 0.33 and 0.67 were categorized as heterozygous inheritance from the mother, whereas variants with BAFs at 0 or 1 were categorized as homozygous inheritance from the mother.
3. To analyze inheritance patterns of triploid-informing-variants, the null hypothesis that inheritance of these maternal variants would be heterozygous was used, arbitrarily, as a starting point. To determine which genomic locations deviated from expected heterozygous inheritance, all BAFs were first normalised to either be 0.33 for heterozygous sites or 0 for homozygous sites. For each triploid-informing-variant, 0.33 was subtracted from the BAF to quantify the deviation from expected heterozygosity (delta heterozygosity).
4. The mean delta heterozygosity was then calculated in 1Mb bins and plotted in Figure 3A on a per chromosome basis.
5. The retention of maternal centromeric homozygosity, with distal recombination, supports that the extra set of chromosomes in the triploid lineage of the proband originates from the same meiosis as the oocyte, which gave rise to the diploid lineage of the patient. This is only consistent with incorporation of the second polar body, when considering alternate events around fertilisation that could lead to forms of triploidy (See explanatory figure below, on next page).

*Alternate hypothesis #1 – Unreduced female gamete.* If the triploidy originated from an unreduced maternal gamete, the proband should have 100% triploidy, which is not the case, and full triploidy is not viable.

*Alternate hypothesis #2 – fusion of two embryos or additional gametes*. Alternate possibilities related to the fusion of two embryos or a second oocyte, would not lead to retention of maternal centromeric homozygosity (with each centromeric haplotype segregating randomly, the odds of fusing two meiotic products from the same person with identical centromeric haplotypes is (0.5)^23^ or p = 1.2^e-7^).

*Alternate hypothesis #3 – incorporation of the first polar body.* Incorporation of the first polar body would lead to retention of 100% maternal centromeric heterozygosity, which is not the case.

*Alternate hypothesis #4 – dispermic origin.* This is not supported by the 43,337 variants coming from the mother and 128 from the father when comparing triploid LCLs *versus* diploid fibroblasts for the patient.

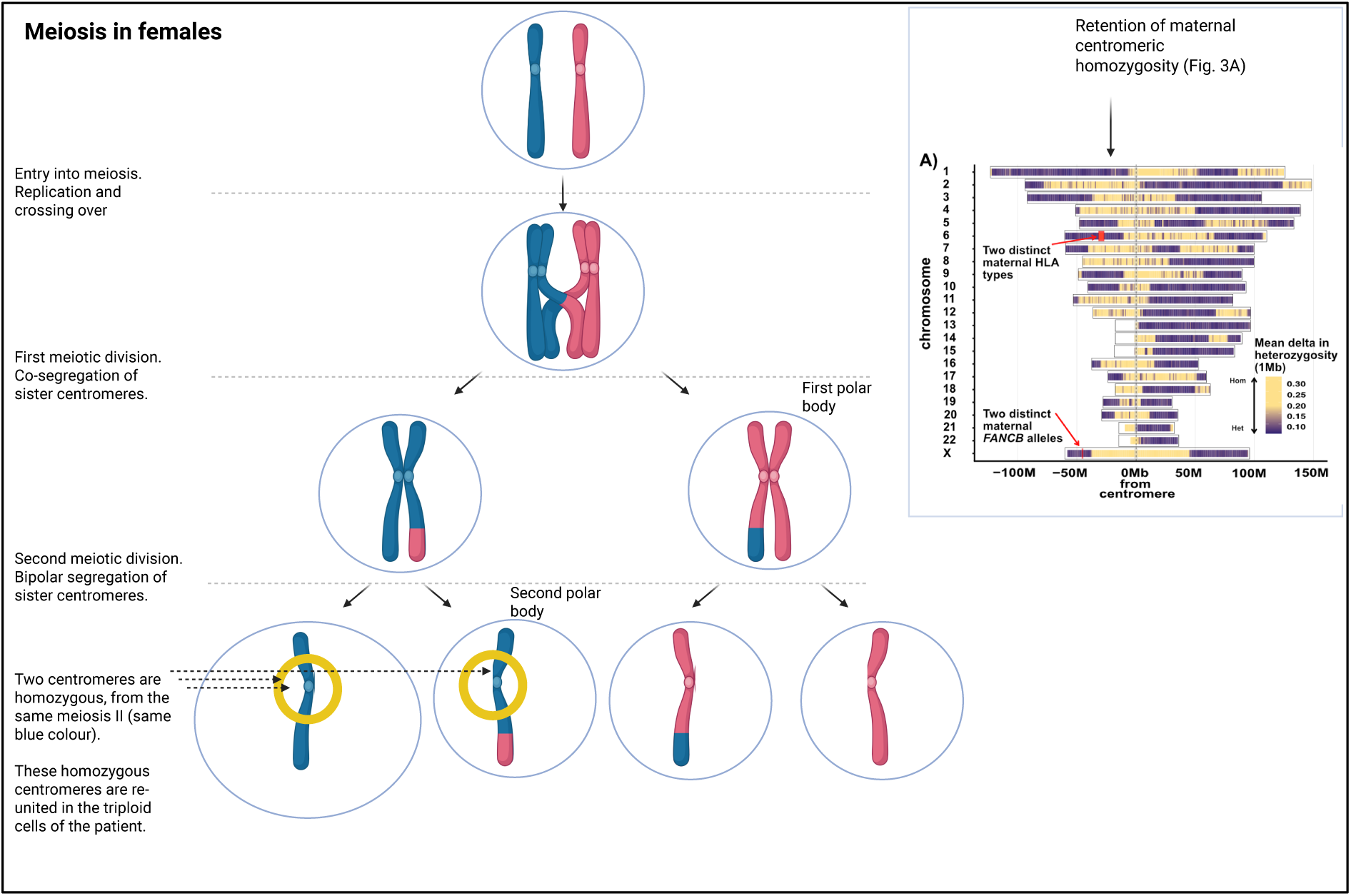

*Differing ploidy across samples (Fig. 3C) –* We next sought to quantify the proportion of cells which were triploid, based on genomic data from different samples. We performed the following analysis:

1. We started with the same 43,337 variants as above, which differ between the patients LCLs and fibroblasts, where the variants were heterozygous in the mother’s germline and homozygous alternate allele in the father’s germline.
2. We filtered for variants which were homozygous alternative in the genome of the proband’s fibroblasts, which results in a median value of approximately BAF = 1.0 in a diploid sample.
3. Plot those variants for all samples, in Fig. 3B.
4. In the case of samples which have any degree of triploidy, the sample median BAF moves towards 0.67 in a sample which is 100% triploid. As the samples can be a mixture of diploid and triploid, values are observed between 0.67 and 1.0.

## Methods

### Ethics

This study was approved by the Royal Children’s Hospital Human Research Ethics Committee. All study participants provided written informed consent. To protect participants’ privacy, medical history is largely omitted. Further, time points of sample collection, ages and dates have been rounded off, and similarly, other information such as the pedigree and pathogenic germline variant segregation data is omitted. Readers can contact the corresponding author to request access to these and other data.

#### Lenti-viral transduction

*FANCB* cDNA was synthesized by Gene Universal and cloned into pAIP using In-Fusion method (Takara). The pAIP plasmid was made by the Luban laboratory^17^. pAIP-FANCB vector was used to generate lentivirus. 500 ng of pAIP-FANCB, 125 ng of VSV-g and 375 ng psPAX2 were mixed in OptiMEM. FuGENE® HD was used as the transfection agent at a ratio of 3:1. Transfection particles were left to form over 20 mins, then were added to HEK293T cells in a well of a six-well tissue culture plate. Virus was collected at 48 and 72h, then pooled, filtered and aliquoted. 1ml of virus was added to hTERT immortalised patient fibroblasts derived from the left side of the body with polybrene at a concentration of 8 µg/mL. The patient’s fibroblasts underwent puromycin selection at a concentration of 0.5 µg/mL. Once selection was complete, these fibroblasts were characterised with the chromosome fragility test, FANCD2 foci assay and mitomycin C survival assays to demonstrate *FANCB* complementation.

### Ploidy analysis of buccal epithelial cells

#### Anatomical pathology – cell type composition

Collection of buccal epithelial cells was performed with using Rovers Orcellex Brushes fixed in a BD Surepath vial. 1 mL of sample was pelleted at 900 x g for 3 minutes. The supernatant was discarded, and the sample was resuspended in 100 µL of sample fixative. Positively charged slides were cleaned with ethanol and samples were adhered using a Shandon Cytospin 4 cytocentrifuge (Thermo Fisher Scientific) at 800 × g for 3 minutes, using the medium acceleration setting. Similarly, cell blocks were prepared by pelleting 1 mL of sample with 400 x g for 7 minutes. Both cytospin and cell block samples were analysed by a trained pathologist and assessed for cell type composition.

#### Cytogenetics – FISH

The buccal epithelial cells – using aliquots of the samples referenced above – were washed once with PBS followed by two rounds of fixative exchange and resuspended in Carnoy’s fixative (three parts of methanol and one part of acidic acid). FISH slides made with the cell suspension were digested using Digest-All (ThermoFisher Scientific catalog number 003009) for 15 minutes at 37°C. The XCE X/Y FISH probe was then added onto the FISH slides followed by a 5-minute co-denaturation at 85°C and overnight hybridisation at 37°C. After the hybridisation, the slides were first washed with 0.4xSSC/0.3% NP40 at 76°C then with 2xSSC/0.1% NP40 at room temperature. 20µL of 3ng/mL DAPI was added onto each slide before the coverslip.

X/Y probe: XCE X/Y XCyting Centromere Enumeration Probe (metasystem: D-0825-200- OG). https://metasystems-probes.com/en/probes/xce/d-0825-200-og/ FISH images were acquired with a Zeiss Axiovision Z2 microscope with Metasystems software Isis v5.10.134.

### FISH slide preparation from urine-derived cells

Urine samples were processed on ice. Cellular material was pelleted by centrifugation at 600 × g for 5 minutes. Pellets containing epithelial-like cells were resuspended in Carnoy’s fixative (3:1 methanol:glacial acetic acid) and incubated at 4°C for 20 minutes. Samples were washed once in fresh Carnoy’s fixative, resuspended in 1 mL fixative, and used for slide preparation. Slides were prepared using a Shandon Cytospin 4 cytocentrifuge (Thermo Fisher Scientific) at 600 × g for 7 minutes and air-dried prior to fluorescence in situ hybridisation (FISH).

### Chromosome fragility assays

Diagnostic chromosome fragility testing was performed in a NATA-accredited laboratory (Pathology Queensland). Additional chromosome fragility assays were performed as previously described^18^. Briefly, patient LCLs were seeded at 250,000 cells/mL in 4 mL of volumes. After 24 h, they were treated with diepoxybutane with 4 different concentrations – 0 µg/mL, 0.01 µg/mL, 0.5 µg/mL and 0.1 µg/mL and left for 70 hours. For fibroblasts, a T75 flasks in log phase was used for each genotype and drug concentration (0 µg/mL, 0.025 µg/mL, 0.05 µg/mL). The cells were harvested by centrifugation at 800 g for 8 mins. Cells were prepared for metaphase spreads by swelling in a hypotonic solution of potassium chloride for 25 mins at 37°C. They were then fixed with fresh-chilled Carnoy’s fixative (3:1 ratio of methanol and acetic acid) for 30 mins. Fixative was removed by centrifugation. Cell suspension was dropped from a height of 25 cm onto a polysine-charged glass slide and left to dry in humidity chamber. Cells were then stained with Giemsa stain and coverslips were mounted onto slides with DPX mounting medium. Metaphases were imaged using a Zeiss Axiovision Z2 microscope with Metasystems software Isis v5.10.134. Images of metaphase spreads were blinded for scoring. Chromosomes were counted to determine ploidy, then chromatid breaks, and chromosome figures were counted.

### FANCD2 foci immunofluorescence

The non-complemented and FANCB-complemented patient-derived fibroblast cells were seeded on sterile glass coverslips at a confluency of 1.5 × 10⁵ cells per well. The next day, cells were treated with 1 μM mitomycin C (MMC) for 5 hours. After washing with PBS, cells were pre-extracted on ice for 8 minutes using pre-extraction buffer (10 mM Tris-HCl pH 7.4, 2.5 mM MgCl₂, 0.5% NP-40, 1 mM PMSF) and washed with ice-cold PBS for a further 3 minutes. Cells were then fixed in 4% paraformaldehyde (PFA) for 15 minutes at room temperature. Blocking was performed using 10% goat serum in 0.1% PBS-Tween 20.

Primary antibody incubation was carried out overnight at 4 °C in a humidified chamber using anti-FANCD2 antibody (cat. #ab108928) at a dilution of 1:200. The following day, coverslips were washed with 0.1% PBS-Tween 20 and incubated at room temperature for 1 hour with Alexa Fluor 488-conjugated secondary antibody (cat. #A32790) at a dilution of 1:300. After washing with PBS-Tween 20, nuclear staining was performed using 300 nM DAPI in PBS for 15 minutes at room temperature. Coverslips were then washed thoroughly in PBS, air-dried, and mounted using ProLong Diamond Antifade. Slides were imaged using Leica Thunder microscope under 63X oil immersion objective with LASX (Leica) software. FANCD2 foci were quantified using CellProfiler software, and results were statistically analysed and plotted using GraphPad Prism.

### Cell survival assays

Patient fibroblasts and the FANCB-complemented fibroblasts were seeded at a density of 5,000 cells per well in a 96-well plate. Cells were then treated with varying concentrations of Mitomycin C (MMC). The growth curve of cells under different MMC concentrations was monitored using the Incucyte system and plotted at 12-hour intervals over 5 days. Growth curves generated over time was based on the cell density in each well. At the endpoint, sulforhodamine B (SRB) staining was performed to assess overall survival after 5 days of MMC treatment.

For the SRB assay, media was removed completely, and cells were fixed with cold 10% trichloroacetic acid (TCAA) for 1 hour at 4 °C. After discarding the TCAA, cells were washed twice with tap water and allowed to air dry completely. Cells were then stained with SRB dye (0.4% SRB in 0.1% acetic acid) for 30 minutes at room temperature. The dye was discarded, and each well was washed twice with 1% acetic acid to ensure no residual dye remained in the wells. Plates were then left to dry overnight at room temperature. The following day, 100 µL of 10 mM Tris was added to each well and incubated at room temperature for 10 minutes. Absorbance was measured at 550 nm using an EnSpire Reader and normalized percentage survival was calculated and plotted for the different MMC concentrations using GraphPad Prism.

### Cytogenetics and ploidy analysis

Karyotyping was performed on bone marrow aspirates and as part of clinical care in NATA accredited laboratories. The bone marrow aspirate samples were cultured in RPMI 1640 medium with 20% FBS for 24 hours before synchronisation with 14 hours of FdU treatment and 7 hours of BrdU treatment. The cells were harvested automatically using Hanabi-PIII Metaphase Chromosome Harvester with 30 minutes of colcemid exposure and resuspended in Carnoy’s fixative (three parts of methanol and one part of acidic acid). Standard cytogenetic techniques were used for slide making and G-banding. Analysis was performed using the Metasystem Metafer AutoCapt scanning sytem and Ikaros karyotyping software.

G banded cytogenetic analysis was performed with 300-400 band resolution. Ploidy determination was also performed in a research setting with patient-derived LCLs and fibroblasts without G-banding. Further, prenatal SNP array analysis was performed on DNA extracted from cultured amniocytes using an Illumina 300K CytoSNP-12 v2.1 microarray to assess for chromosomal aneuploidies.

### Non-invasive prenatal testing

Non-invasive prenatal testing (NIPT) was performed using the Harmony assay (Australian Clinical Labs, Melbourne). Two maternal plasma samples failed internal quality-control metrics and did not yield reportable results for the chromosomes considered by the test: 13, 18 21. A subsequent plasma sample collected was analyzed at the Victorian Clinical Genetics Services (VCGS, Melbourne) using the percept assay, which applies low-coverage whole-genome sequencing of cell-free DNA^19^. This analysis passed quality control and returned a low risk result for all chromosomes. Triploid mosaicism is not assessed by this assay.

### Genomic sequencing and analyses. Whole-exome sequencing analysis

DNA was extracted from peripheral blood from maternal, paternal, and patients samples. Libraries were prepared for whole-exome sequencing (WES) using an Agilent SureSelect XT Low Input Clinical Research Exome x2 (Agilent CRE v2) kit, and sequenced with an Illumina NovaSeq6000. FASTQ files were aligned to the human reference genome (GRCh38/hg38, Genome Reference Consortium) using BWA-MEM^20^ (Li and Durbin, 2009). Downstream processing of BAM files, including addition of read groups, marking of duplicates, and base quality score recalibration, was performed using the Genome Analysis Toolkit (GATK)^21^, with known variants from dbSNP138 (UCSC Genome Browser). Variant calling was performed using GATK HaplotypeCaller in GVCF mode (-ERC GVCF), restricted to exonic regions using a custom BED file. Joint genotyping across the trio was conducted with GATK GenomicsDBImport and GenotypeGVCFs.

### High molecular weight DNA extractions

High molecular weight genomic DNA was extracted using the Nanobind PanDNA Kit (Circulomics, 103-260-000). Lymphoblastoid cell lines were processed following the manufacturer’s protocol for suspension cultured cells (103-394-500 REV01 FEB2024), while fibroblasts were processed following the protocol for adherent cultured cells (102-573-600 REV03 MAR2024). DNA concentration and purity were assessed using a Qubit Fluorometer (1X High Sensitivity dsDNA assay; Thermo Fisher Scientific) and a Nanodrop spectrophotometer (Thermo Fisher Scientific). DNA integrity was evaluated using the TapeStation 4200 system (Agilent Technologies).

### PacBio HiFi sequencing

High molecular weight DNA quality and fragment size distribution were assessed by the Australian Genome Research Facility (AGRF, Queensland) using a Femto Pulse system (Agilent Technologies). Sequencing was performed by AGRF on a PacBio Revio System (Pacific Biosciences) using one Revio SMRT Cell.

### ONT Long-read library preparation and analysis

Long-read sequencing libraries were prepared using the Ligation Sequencing Kit V14 (SQK- LSK114; Oxford Nanopore Technologies), according to the manufacturer’s instructions. Libraries were loaded onto a PromethION flow cell (FLO-PRO114M) and sequenced on a PromethION 2 Solo instrument (Oxford Nanopore Technologies).

Oxford Nanopore long-read genomic DNA sequencing was performed on lymphoblastoid cell lines (LCLs) and fibroblast samples from the patient. Reads were aligned to the GRCh38/hg38 reference genome using minimap2^22^ with the “-ax map-ont” setting, and aligned files were converted to sorted BAM files using samtools^20^. Variant calling for single nucleotide polymorphisms (SNPs) and short insertions and deletions (INDELs) was conducted using Clair3^23^.

### Short reads

Genomic DNA was submitted to the Australian Genome Research Facility (AGRF) for library preparation and sequencing. Libraries were prepared using the Illumina DNA Prep PCR-Free kit and sequenced on an Illumina NovaSeq 6000 platform, generating 150 bp paired-end reads. Short-read whole-genome sequencing (WGS) at 60x coverage was performed on the patient’s peripheral blood mononuclear cells (PBMCs) and bone marrow (BM) at the most recent collection, as well as peripheral blood from other family members. Reads were aligned to the GRCh38/hg38 reference genome using BWA-MEM, followed by duplicate marking, read group addition, and base recalibration with GATK and dbSNP138. Variant calling was performed using HaplotypeCaller and GenotypeGVCFs.

### Identity by descent analysis

For identity-by-descent (IBD) analysis, VCF files from the patient and obligate female carriers were merged using bcftools merge, then analysed in R to identify variants heterozygous in obligate female carriers but hemizygous in the patient. Variants on chromosome X within the *FANCB* locus were assessed for pathogenicity and searched for in variant databases including ClinVar and gnomad.

### Long-read RNA isoform detection

All cell lines were preserved in RNAlater and stored at −80°C prior to RNA extraction. Total RNA was isolated using the ISOLATE II RNA Mini Kit (Bioline) following the manufacturer’s protocol. RNA purity was assessed using a Nanodrop spectrophotometer, concentration measured with a Qubit Fluorometer (RNA Broad Range Assay; Thermo Fisher Scientific), and RNA integrity evaluated on the TapeStation 4200 (Agilent Technologies).

For long-read RNA sequencing, 500 ng of total RNA per sample was processed using the cDNA-PCR Barcoding Kit (SQK-PCB114.24; Oxford Nanopore Technologies). First-strand cDNA synthesis and strand-switching were performed according to the manufacturer’s protocol. PCR amplification of full-length cDNAs was carried out using LongAmp Taq 2X Master Mix (New England Biolabs) under the following cycling conditions: 95°C for 30 s; 14 cycles of 95°C for 15 s, 62°C for 15 s, and 65°C for 6 min; followed by a final extension at 65°C for 6 min. Barcoded libraries were pooled equimolarly, targeting a total of 50 fmol cDNA (12.5 fmol per sample), based on an average transcript length of ∼3 kb (*FANCB*). Final adapter ligation, purification, and flow cell priming were performed according to the manufacturer’s instructions. Sequencing was performed on a PromethION 2 Solo using a single PromethION flow cell (FLO-PRO114M). Data were collected using MinKNOW software (Oxford Nanopore Technologies). Sequencing data was aligned to GRCh38/hg38 using minimap2 with “-ax splice” for spliced alignment. Control data was downloaded from Sequence Read Archive, accession number ERR13350351. Visualization in Integrative Genomics Viewer (IGV) identified intron retention in the region with *FANCB* variant c.1326+594del identified via IBD analysis.

### Allele frequency analysis

Allele frequency (AF) analysis was performed by genotyping BAM files from LCLs, fibroblasts, PBMCs, bone marrow, and WES samples using bcftools mpileup to include homozygous reference sequences. Informative SNPs were defined as sites with a homozygous paternal genotype and heterozygous maternal genotype. AF calculations were performed in R based on read counts (AD_REF and AD_ALT) and calculated as a proportion of total reads called for each variant. Minor allele frequencies (MAF), calculated from AD_ALT, were used for additional analysis, which indicated that the patient LCLs are triploid with an additional maternal genome. Examination of homozygous and heterozygous site distribution relative to centromeres showed increased homozygosity at centromeres, consistent with retention of the second polar body.

Guassian mixture modelling was conducted on the MAF’s in R to determine the degree of ploidy in each sample. The results demonstrated that the cultured LCLs and the bone marrow samples had a high degree of triploidy. The PBMCs taken at a later stage had a relatively increased triploid fraction. The fibroblasts were largely diploid, as were the PBMCs taken at an earlier stage for WES.

### Oligos used in this study

MS137 5’-GGAACCTTGGGGAACAAGTC-3’

MS138 5’-GGCACTACAGTGACTTGAGG-3’

MS140 5’ - CCCAGGTTCAAGTTATTCTCCT-3’

MS144 5’-GAGAATCGTTACCTGGTGGT-3’

MS145 5’-CACTCCAACAACCAAGCTATC-3’

MS147 5’-ATGAGCCACCACGCCCGG-3’

MS148 5’-CAAGAGATTGAGACAGTCCT-3’

Minigen FANCB Forward-XhoI 5’-CTTCTCGAGTTGAAATCATGTTTCTTGCCACT-3’

Minigen FANCB Reverse-BamHI 5’-GTTAGGATCCGACCCAAACCCTTACATGGC-3’

Forward M1 5’-CTGGCCCTGCTCATCCTCT-3’

Reverse M2 5’-CAGTGCCAAGGTCTGAAGGT-3’

Forward-cDNA: 5’-TTTTCGGGAATTACGGCAGC-3’

Reverse-cDNA: 5’-TCCCTTCGAATGGTGTTCTCT-3’

### Targeted *FANCB* cDNA analysis

Total RNA from RPE cells and fibroblast samples was treated with DNase I (New England Biolabs) to remove genomic DNA. DNase reactions were cleaned using RNAclean XP beads (Beckman Coulter), and RNA was eluted in nuclease-free water. RNA integrity and concentration were confirmed by spectrophotometry prior to cDNA synthesis.

For each sample, 1 µg of total RNA was reverse transcribed using Maxima H Minus Reverse Transcriptase (Thermo Fisher Scientific) and oligo(dT) primers, following the manufacturer’s protocol.

PCR was performed to amplify the poison exon positioned between exons 5 and 6 of *FANCB*. Each 10 μL reaction mixture contained 1 μL of the cDNA template (2:3 diluted cDNA extract: milliQH2O), 2 μL of MyTaq Reaction Buffer Red (Meridian Bioscience, Cat: BIO-37112), 0.5 μM each forward and reverse primer, and 0.1 μL of MyTaq HS DNA polymerase (Meridian Bioscience, Cat: BIO-21112). Cycling conditions were: initial denaturation at 95℃ for 1 minute; 35 cycles of 95°C for 30 s, 55°C for 30 s, and 72°C for 1 minute; and final extension at 72°C for 5 minutes. Three pairs of primers were used to target the poison exon: (1) MS144 and MS145, flanking exon 4-6 of FANCB, yielding a 291 bp amplicon (390 bp if poison exon is present); (2) MS144 and MS147, amplifying region between FANCB exon 4 to poison exon, with MS147 located within the poison exon, yielding a 195 bp amplicon ; (3) MS148, amplifying region between the *FANCB* poison exon to exon 6, with MS148 located within the poison exon, yielding a 176 bp amplicon. Amplicons were visualised in a 2% Agarose gel at 100V for 60 minutes.

### Genotyping of cell lines and patient samples for the *FANCB* variant

To genotype cell lines and patient samples for the *FANCB* variant (NM_001018113.3:c.1326+594del) we amplified the region containing the variant and confirmed its presence with Sanger sequencing at the Australian Genome Research Facility. The oligo pair used in the PCR was MS137 and MS138. MyTaq polymerase was used to amplify the DNA. Amplicons were purified with Monarch® PCR clean up kit (New England Biolabs) Amplicons were sequenced in both directions with the oligos MS137 and MS140.

### Minigene assay

To determine the pathogenic effect of the *FANCB* c.1326+594del variant, a minigene assay was performed. The genomic DNA fragment comprising the variant was amplified with self-designed oligonucleotides which incorporated XhoI (R0146L, New England Biolabs) and BamHI (R0136L, New England Biolabs) restriction sites and the genomic region spanning exon 6 and the intronic sequence of interest was cloned into the Exontrap Cloning Vector pET01 (MoBiTec GmbH, Göttingen, Germany) from a healthy donor positive control (C+) and patient constructs (PAT). The C+ and PAT plasmids were transfected into HEK293T cells using Lipofectamine3000 (Promega, Madison, WI, USA) following the manufacturer’s protocol. Total RNA was extracted 48 hours after transfection, reverse-transcribed, and amplified by PCR using primers corresponding to the 5 ′ and 3 ′ pET01 exons (M1 and M2, respectively). HEK293T cells transfected with the C+ plasmid showed a single product of expected size on agarose gel, consistent with normal splicing. In contrast, cells transfected with the PAT construct displayed a band of higher molecular weight, indicating the presence of retained intronic sequence. Sanger sequencing of the RT-PCR product confirmed the retention of a 110 bp intronic fragment, matching exactly the aberrant transcript identified in the patient. These results provide strong evidence that the c.1326+594del variant is responsible for FANCB cryptic splice site activation and aberrant RNA processing, supporting its pathogenicity.

### Statistical methods

Supplementary information about statistical testing of cellular assays and data related to Figure 2

Chromosome fragility was quantified by counting aberrant cells per sample. Statistical comparisons between relevant cell lines and DEB concentrations were performed using Fisher’s exact test on the number of aberrant cells. To account for the zero counts in some groups, we applied the Haldane–Anscombe correction, which adds 0.5 to all cells in a contingency table, allowing the calculation of odds ratios without introducing bias. For visual representation in the main text, we plotted the number of chromosomal aberrations per cell to illustrate the extent of chromosomal damage in Figures 2A, 2D and 2G.

### Chromosome fragility tests (Figures 2A, 2D, 2G)

**Supplementary table 1.** Corresponding to Figure 2A. Complementation with *FANCB* rescues the chromosome fragility phenotype of the patient’s immortalized left-side fibroblasts at the highest DEB concentration tested (0.05 µg/mL; Fisher’s exact test, Haldane-Anscombe-corrected odds ratio 70, 95% CI 10–500, p < 0.001).

| DEB concentration $\mu\text{g/mL}$ | Aberrant cells - patient left fibroblasts | Aberrant cells - patient left fibroblasts + *FANCB* | Odds ratio | 95% CI | p-value |
| --- | --- | --- | --- | --- | --- |
| 0 | 8/25 (32%) | 1/25 (4%) | 10.8 | 1.3–52 | 0.023 |
| 0.025 | 4/25 (16%) | 1/25 (4%) | 4.4 | 0.4–23 | 0.35 |
| 0.05 | 21/25 (84%) | 1/25 (4%) | 70 | 10–500 | <0.001 |

**Supplementary table 2.** Corresponding to Figure 2D. Patient LCLs do not have a chromosome fragility phenotype, when considered as one population of cells, and are not different from non-FA LCLs at the highest DEB concentration tested (0.1 µg/mL; Fisher’s exact test, Haldane-Anscombe-corrected odds ratio 0.8, 95% CI 0.2–3.3, p > 0.99).

| DEB concentration µg/mL | Aberrant cells - Patient LCLs | Aberrant cells - LCL608 (non-FA) | Odds ratio | 95% CI | p-value |
| --- | --- | --- | --- | --- | --- |
| 0.0 | 0/25 (0%) | 2/25 (8%) | 0.2 | 0.0-4.0 | 0.49 |
| 0.1 | 3/50 (6%) | 4/50 (8%) | 0.8 | 0.2-3.3 | 1.00 |

**Supplementary table 3.** Corresponding to Figure 2G. Patient LCLs contain two populations of cells, diploid and triploid, and the diploid and triploid cells have different chromosome fragility phenotypes at the highest DEB concentration tested (0.05 µg/mL; Fisher’s exact test, Haldane-Anscombe-corrected odds ratio 51.1, 95% CI 2.6-1015, p < 0.001).

| DEB concentration µg/mL | Aberrant cells – Patient diploid LCLs | Aberrant cells – Patient triploid LCLs | Odds ratio | 95% CI | p-value |
| --- | --- | --- | --- | --- | --- |
| 0.00 | 4 (4.12%) | 1 (1.67%) | 1.9 | 0.3-12.5 | 6.50e-01 |
| 0.01 | 11 (29.73%) | 0 (0%) | 66.4 | 3.8-1166 | 1.47e-06 |
| 0.05 | 5 (38.46%) | 0 (0%) | 51.1 | 2.6-1015 | 4.95e-04 |

### Mitomycin C sensitivity (Figure 2C)

Each IC50 value represents a technical replicate of a mitomycin C survival assay, from two independently made *FANCB* lenti-virus transductions. Given the small number of replicates, some comparisons did not reach statistical significance despite consistent biological effects observed with complementary assays in Figures 2A-C. For these comparisons, Welch’s t-test, which does not assume equal variances, was used, and exact p-values are reported for transparency.

---

**Supplementary Figure 1.**
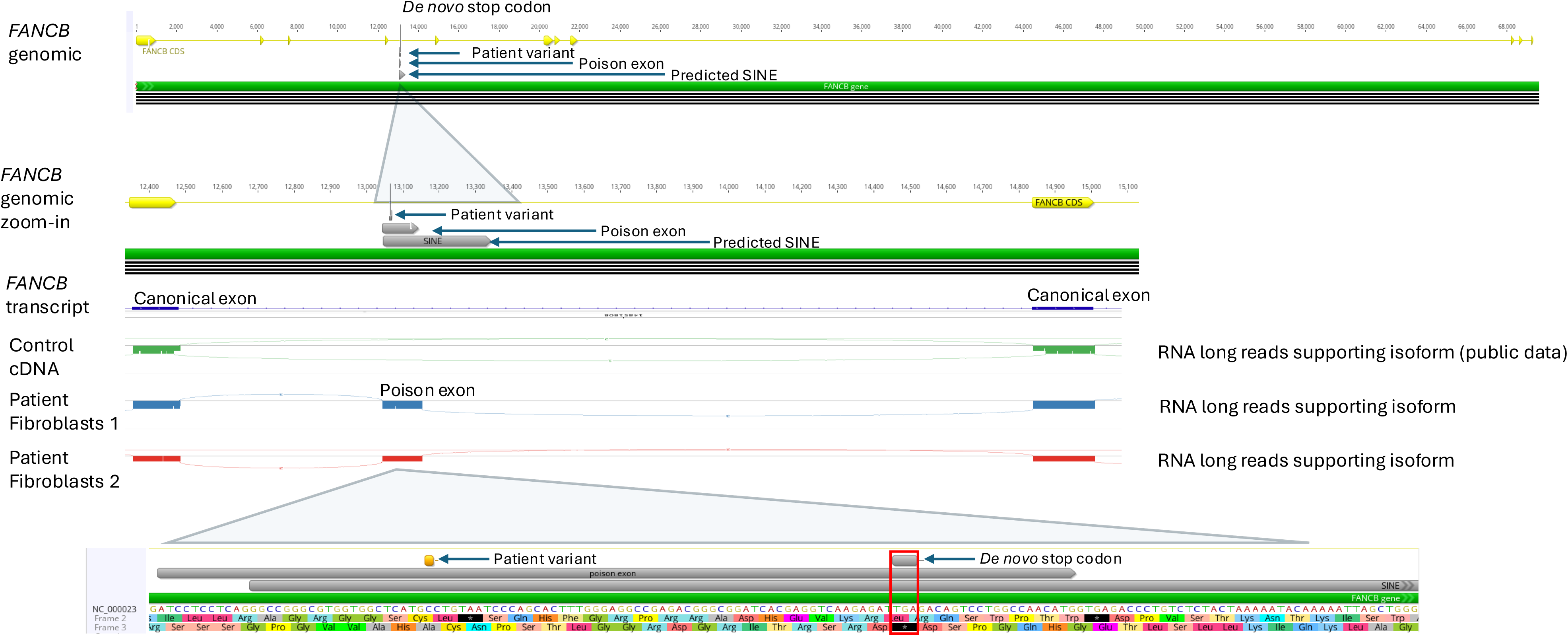
Long-read RNA sequencing supports the retention of an additional 110 nucleotides in the *FANCB* transcript. The additional transcribed nucleotides are associated with a single nucleotide deletion, *FANCB* (NM_001018113.3:c.1326+594del), that creates a poison exon with a premature stop codon. Independent patient fibroblasts support the inclusion of this sequence into the *FANCB* transcript and coverage is absent from public databases.

**Supplementary Figure 2.**
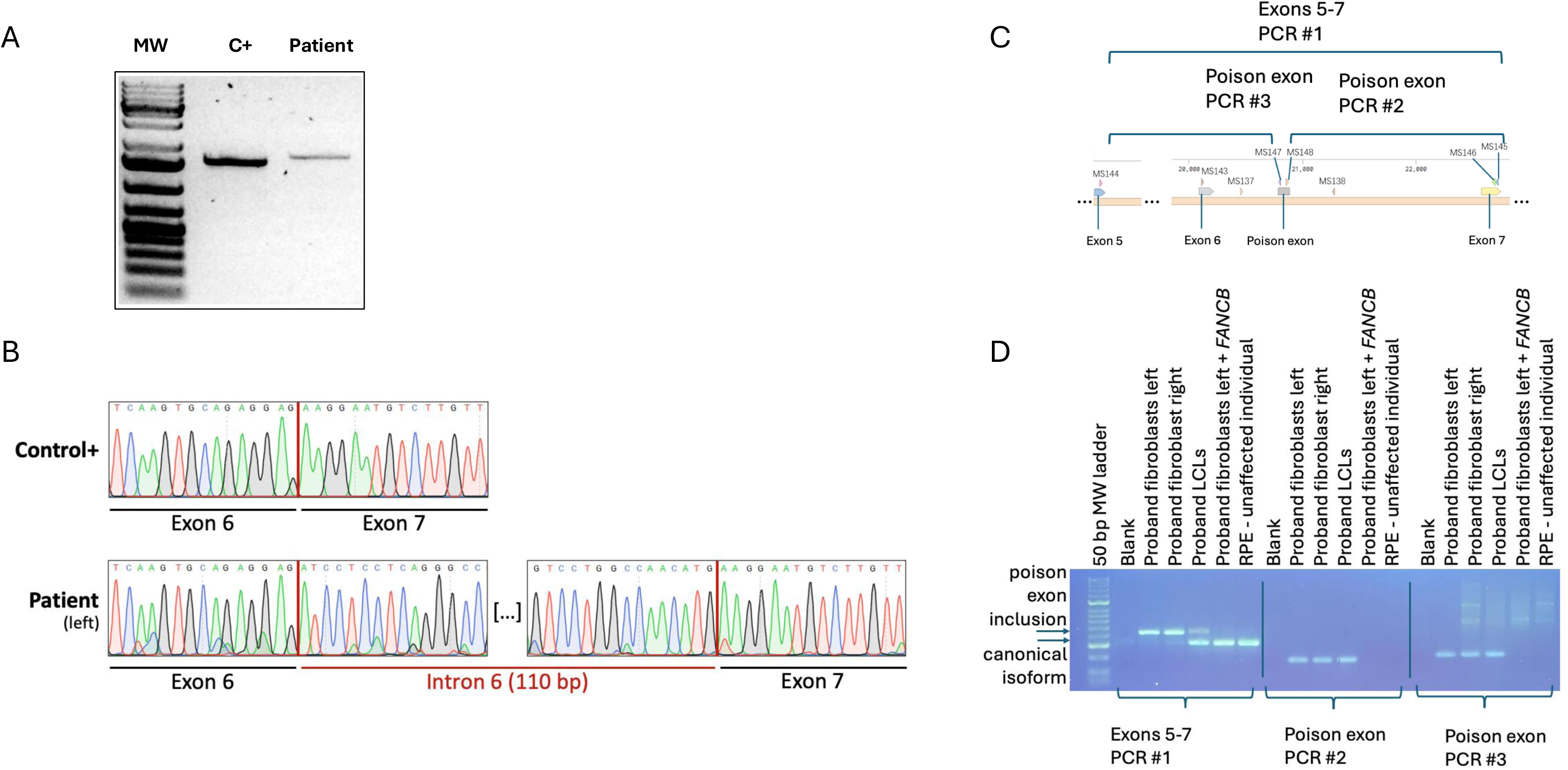
*FANCB* cDNA analysis supports the inclusion of a poison exon in patient fibroblasts and LCLs. Independent patient fibroblasts and LCLs support the exonization of a portion of transposable element into the FANCB transcript. A) cDNA PCRs of control and patient samples for Sanger sequencing. B) Sanger sequencing support the retention of a poison exon. C) cDNA PCR strategy to detect the poison exon in different sample types. D) PCRs on patient and control samples. Both maternal isoforms of *FANCB* are detected in LCLs.

**Supplementary Figure 3.**
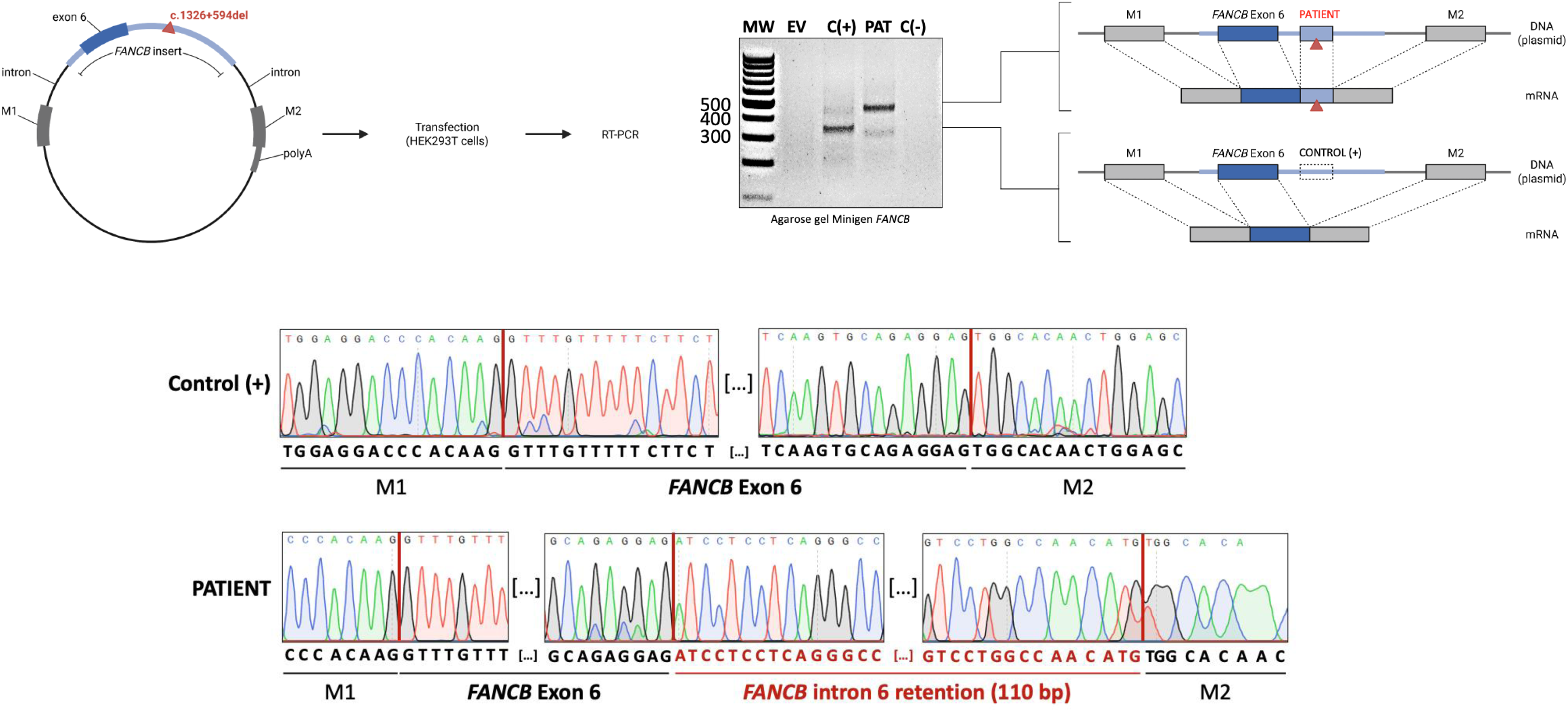
*FANCB* (NM_001018113.3:c.1326+594del) causes poison exon retention in a minigene assay. A) Patient or control DNA from FANCB was cloned into pET01 and transfected into HEK293T cells. B) cDNA was synthesized and amplified . C) Sanger sequence analysis revealed poison exon retention in the sample with the patient *FANCB* variant.

**Supplementary Figure 4.**
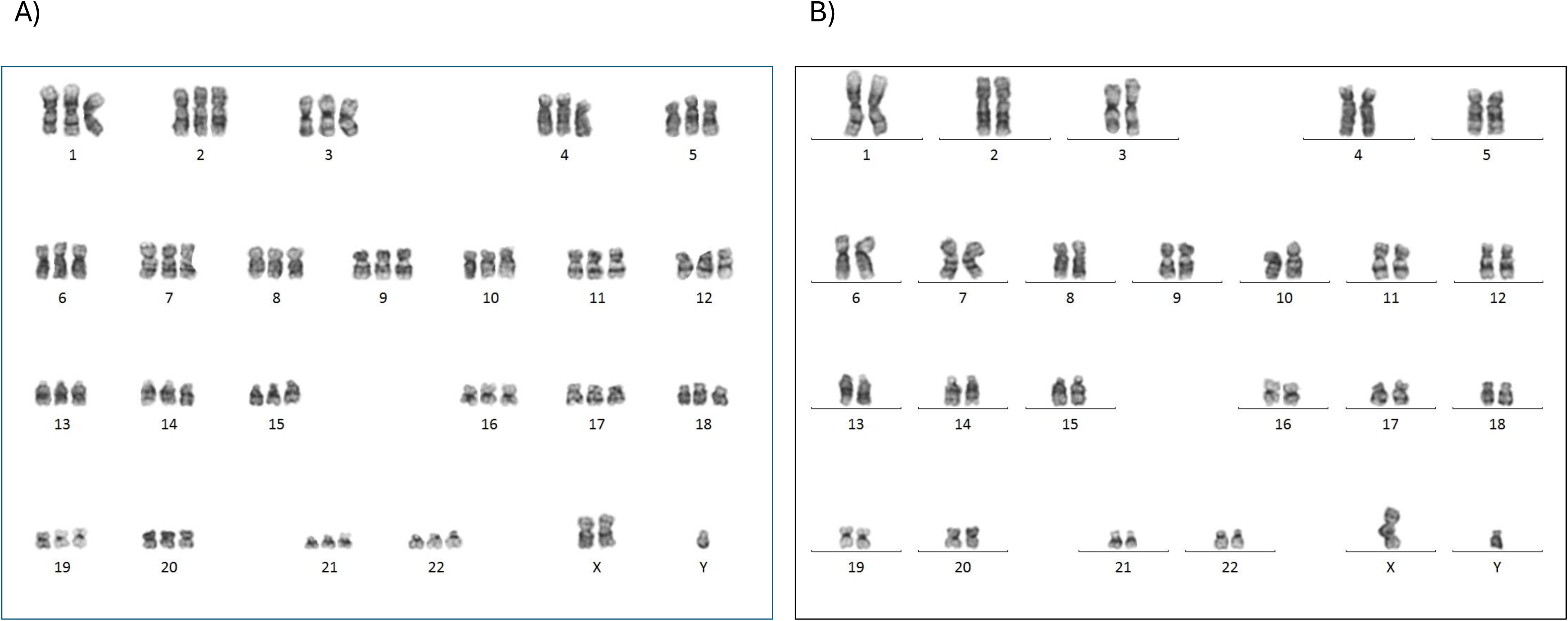
Bone marrow aspirate reveals triploid-diploid mosaicism. Cytogenetics using G-banding on metaphase preparations from bone marrow revealed cells with karyotypes A) 69,XXY or B) 46,XY.

**Supplementary Figure 5.**
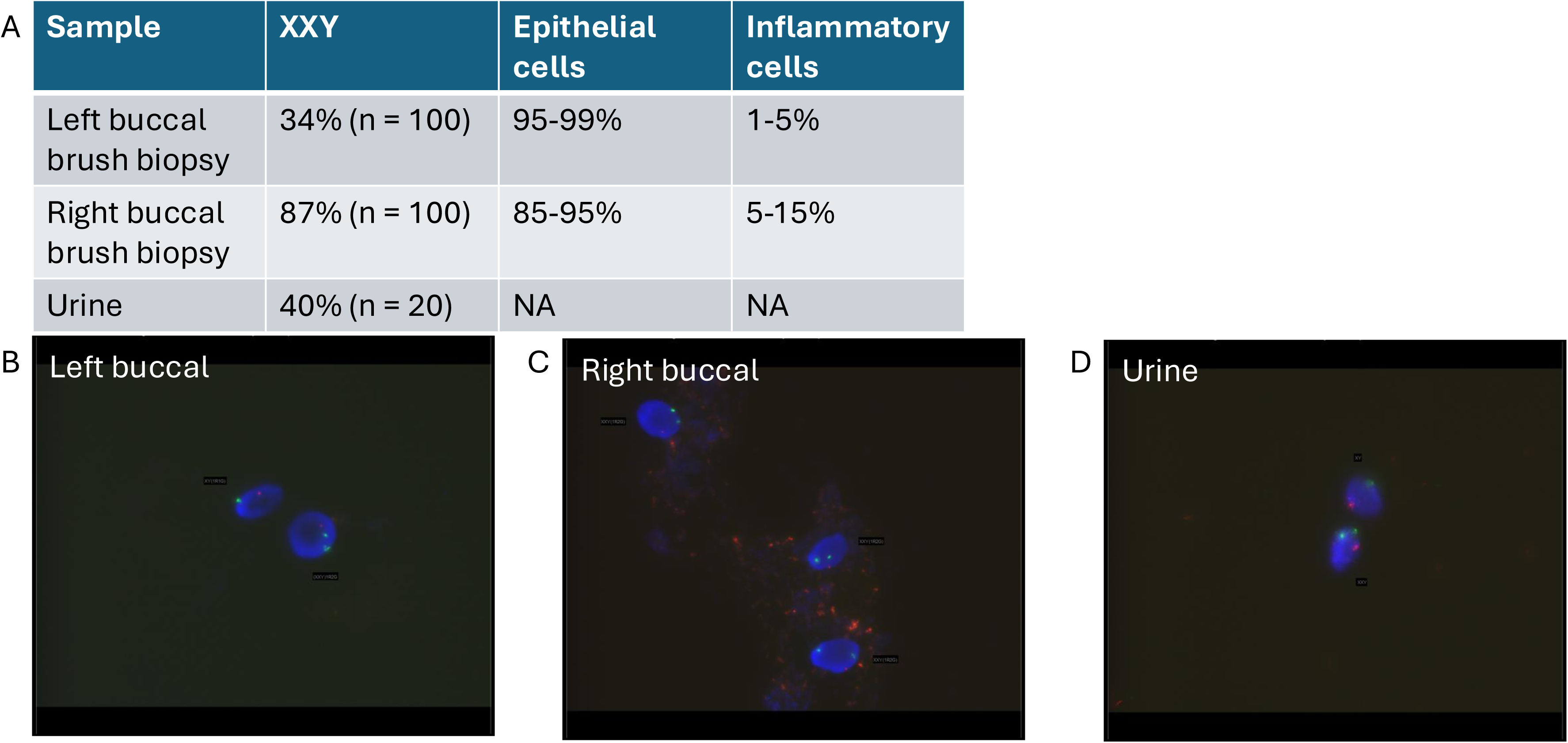
Buccal cell and urine FISH supports germline mosaicism. A) FISH analysis for the quantification of ploidy was coupled with cell type analysis. Epithelial and inflammatory cells are presented as a range because each sample was prepared twice, and each was analysed by a trained pathologist. Given that the proportion of epithelial cells is 85-99%, at least a proportion of the triploid cells must be epithelial. B) Representative FISH images are shown for the left buccal Orcellex brush biopsy, C) right buccal Orcellex brush biopsy, and D) cells isolated from urine. The X and Y chromosome probes are green and red respectively.

**Supplementary Figure 6.**
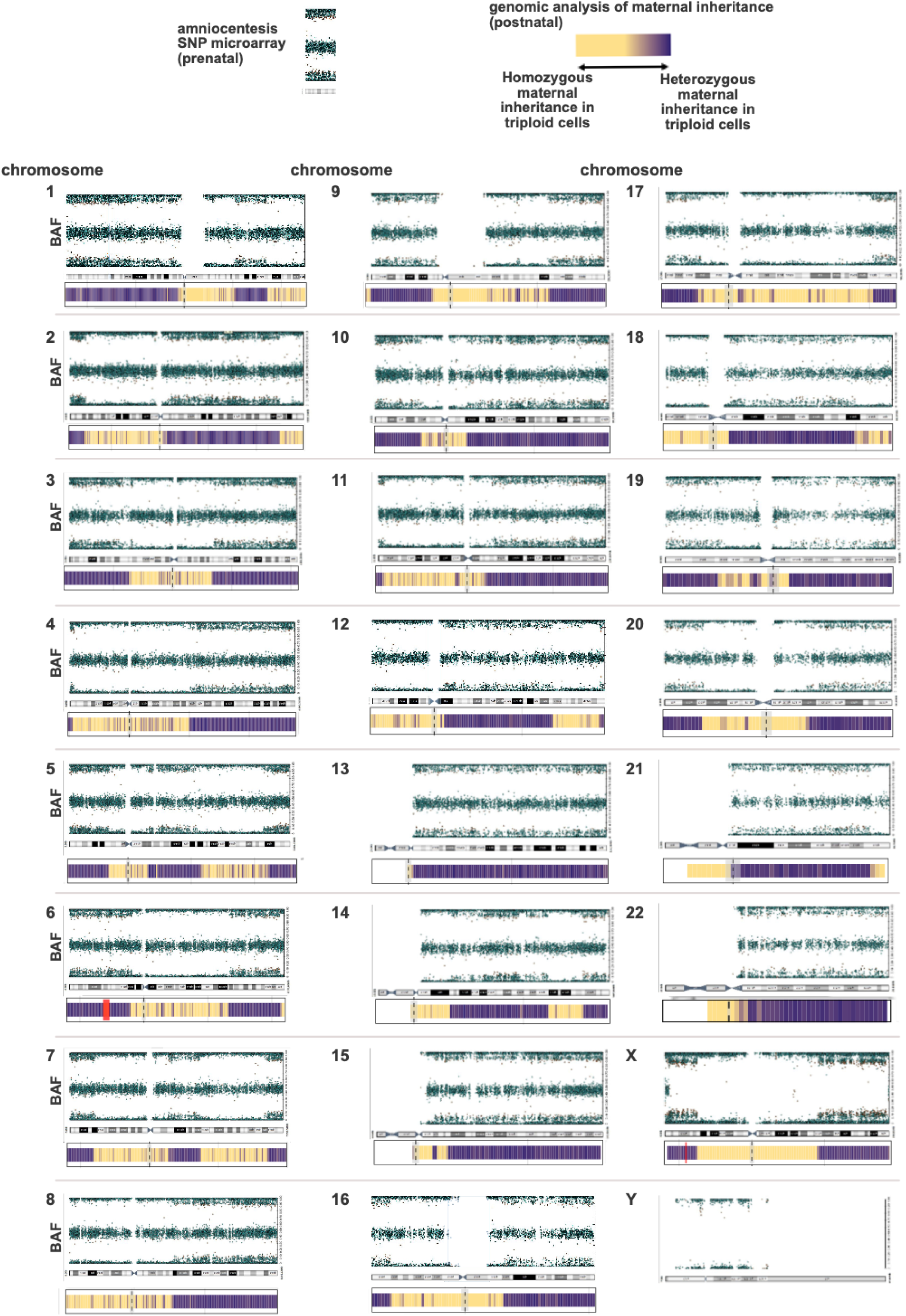
Triploid embryonic mosaicism can be detected in prenatal and postnatal samples. A comparison of postnatal and prenatal sample that both detect concordant embryonic triploid mosaicism profiles. For the postnatal samples, genomic analysis from Figure 3A discriminates between heterozygous or homozygous maternal inheritance in the patient’s triploid cells (lower rows). For the the prenatal sample, amniocentesis SNP microarray data (upper rows) is shown. Amniocentesis B allele frequencies (BAF) are shown. When the BAF = 0.0, the genotypes are AA(mat1pat1) + AAB(mat1,pat1,mat2) in blocks where the additional allele is being introduced from the low grade mosaic triploid cell lineages (the additional allele from recombination at mat M1). Where the BAF is 0.0 (around centromere), the genotypes are AA +AAA). The opposite is true near BAF = 1.0 (BB +BBA) and at BAF = 1.0 (BB +BBB). The two data types in the figure are aligned manually to highlight the concordance between recombination events. Chromosome physical lengths are not to scale. The red highlighted region on chromosome 6 correspond to the *HLA* locus. The red highlighted region on the X chromosome corresponds to the *FANCB* locus. Small acrocentric chromosomes 13, 14, 15, 21 and 22 have regions on their short arms where there is “no data” which appear white. This is because the regions are highly repetitive, and variants cannot be called from the genomic data or probes are not designed for these regions on the SNP microarray. There is a discrepancy in the boundaries of “no data” for the short arms of chromosomes 21 and 22, because the long read data could nonetheless call variants in a subset of these repetitive regions. The heterozygous (BAF ∼0.5) on terminal Xp represents PAR1 with approximately 2.7 Mb of homologous DNA mapping to terminal Xp and Yp. By convention, the array data only presents the cross hybridization for chromosome X.

**Supplementary Figure 7.**
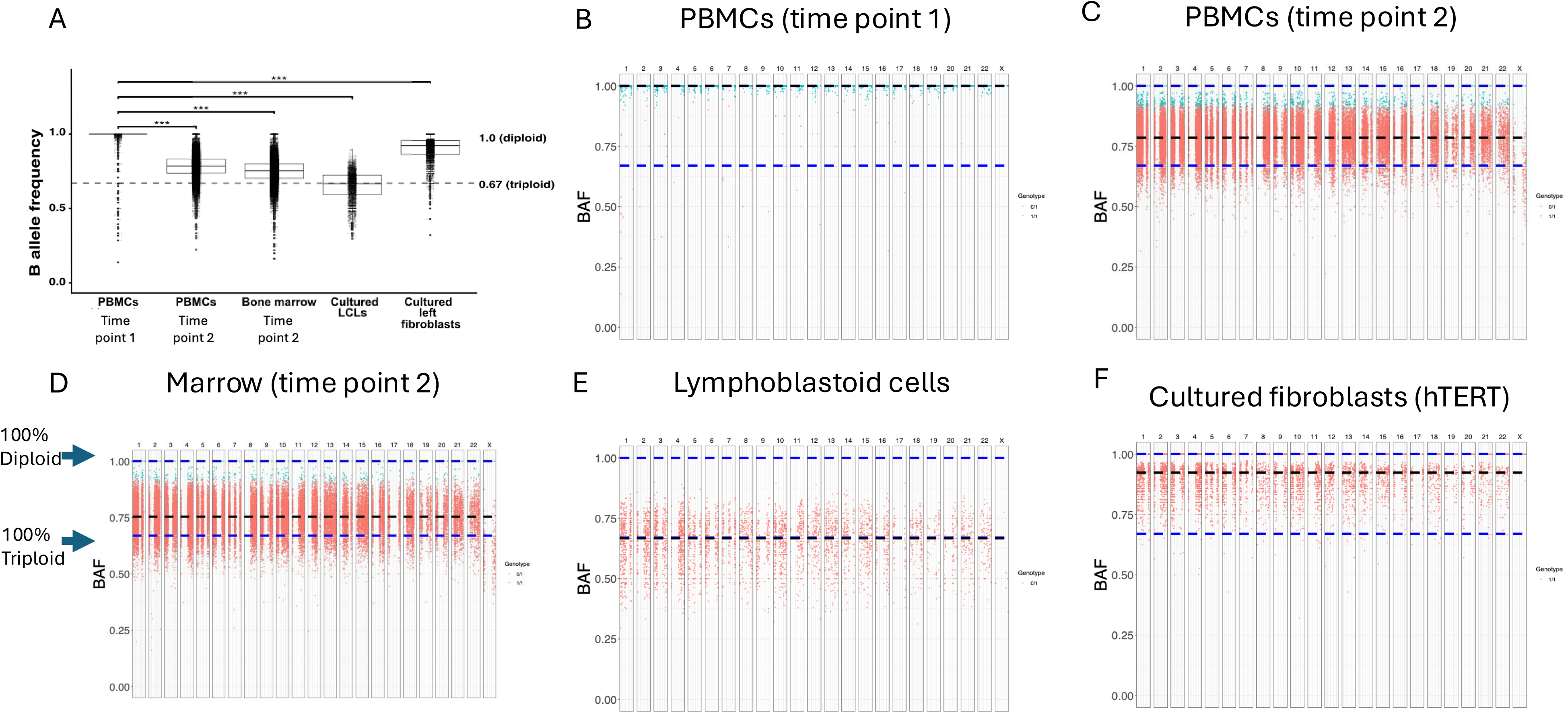
Progressive genome-wide expansion of a triploid hematopoietic lineage. A) Average genome wide readout of ploidy. Individual chromosomes for: B) Whole exome sequencing of peripheral blood at time point 1, and C) time point 2. D) Bone marrow time point 2, E) cultured lymphoblastoid cells show a predominant triploid lineage based on variant allele frequencies of “triploid defining” variant, and F) cultured immortalised fibroblasts. The black dotted line is the median BAF.

**Supplementary Figure 8.**
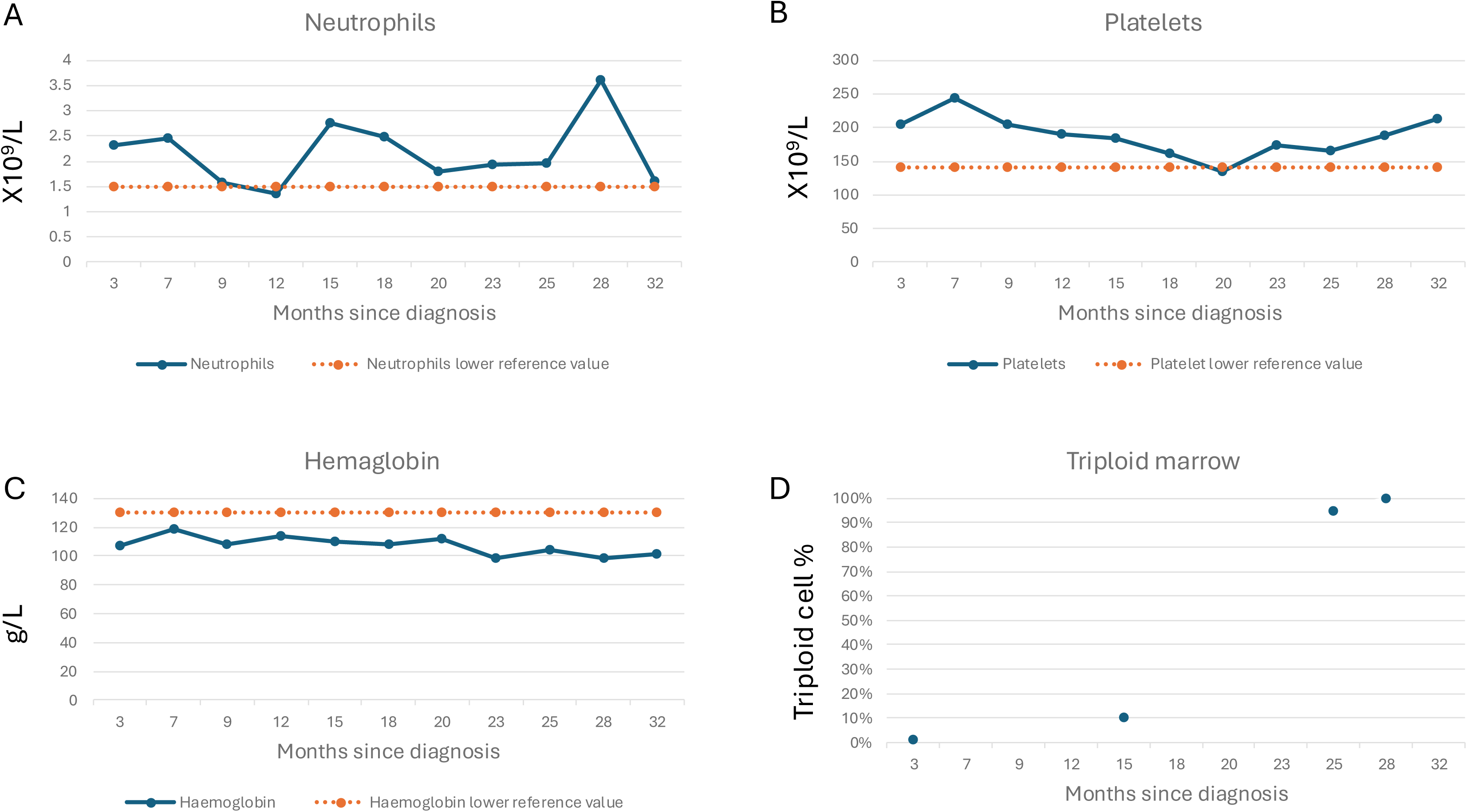
Longitudinal complete blood counts since diagnosis (months). A) Neutrophils, B) platelets, C) hemaglobin, and D) the percentage of cells which were triploid (69,XXY) based on karyotyping of bone marrow samples (3 months post diagnosis, 1%; 2/169, 15 months, 10%; 11/110, 25 months 95%; 104/110, 28 months 100%; 215/215).

